# Mosquito repellent efficacy of Australian Blue Cypress *Callitris intratropica* essential oil and its topical formulation under laboratory and field conditions

**DOI:** 10.1101/2023.02.01.23285221

**Authors:** Melanie Koinari, Brogan Amos, Michael Townsend, Stephan Karl

## Abstract

Mosquito repellents are important for personal protection against nuisance and potentially infectious mosquito bites. Repellent activity of Australian Blue Cypress (ABC) essential oil and a commercially formulated skin lotion containing ABC oil were compared with 20% DEET (N, N-diethyl-3methylbenzamide) and evaluated against mosquitoes under laboratory and field conditions in North Queensland, Australia. Using the arm-in-cage method, the following cumulative doses of ABC oil in ethanol were tested; 0.5%, 1.5%, 3%, 5%, 7.5% and 10.5% against female *Aedes aegypti*. In the field, 2.5%, 5% and 10% of diluted ethanolic ABC oil were tested using the human landing catch method. We observed a dose-dependent increase in protection with diluted ABC oil approaching 80% at high concentrations. While some protection was afforded, mosquito landing and probing was still recorded immediately after application (laboratory) for both ABC essential oil and skin lotion. Protection declined from 80 – 70% and 93 – 50% for 20% ABC oil (laboratory) and 10% ABC oil (highest dose, field), respectively. For the formulated product, protection declined from 85 – 75% in the laboratory and from 63 – 50% in the field. To conclude, both ABC essential oil and the formulated skin lotion provided moderate protection against mosquito bites, which decreased soon after application.

## Introduction

Topical repellents are a key personal protection measure when other methods of protections such as bed nets or area repellents (e.g., mosquito coils) are not possible or practical [1]. Personal protection can influence the infection rates and reduction of mosquito-borne diseases, for example, as observed with malaria [1]. Mosquitoes transmit malaria, filariasis, zika, dengue, and other febrile illness and arboviral encephalitis [2-4]. In Australia, especially, the *Aedes* and *Culex* mosquitoes are known to transmit a number of arboviruses including Ross river, Barmah forest, dengue, Murray Valley encephalitis, Kunjin and Japanese encephalitis [5, 6]. Mosquitoes use a combination of olfactory, visual and thermal cues to detect suitable hosts [7-11]. Mosquitoes use a combination of olfactory, visual and thermal cues to detect and orient towards suitable hosts [7-11]. A repellent is defined as any compound that has an effect on the behaviour of mosquitoes including ‘movement away from the source’ (repellency in the strict sense), as well as ‘inhibition of attraction’ (interference with host detection and/or feeding response) [12, 13].

The use of insect repellents dates back to ancient times, when substances such as plant oils, smokes and tars were used to kill or repel insects [14, 15]. Citronella oil, dimethyl phythalate, Indalone® and Rutgers 612 were the common insect repellents prior to World War 2 [15]. But they failed to provide the desired protection for military personnel, which led to the screening of 20, 000 potential mosquito repellent compounds by the United States (US) government and the discovery of N, N-diethyl-m-toluamide (DEET) [15]. DEET was first introduced into the market in 1956 and serves as an effective broad-spectrum insect repellent with a long-lasting effect on mosquitoes, ticks, chiggers and fleas [15, 16]. DEET is available in a variety of formulations including aerosols, creams, lotions, sprays and in concentration ranging from 5% to 100%, although most products contain less than 40% [16]. Before they can be registered, most skin-applied repellents are evaluated for their safety and efficacy by the US Environmental Protection Agency (EPA) [17]. The US EPA completed re-registration for DEET in 1998 [18]. Apart from DEET, only few other synthetic active ingredients (picaridin and IR3535) are EPA registered for skin-application [17]. Currently, para-menthane 3, 8 diol, (PMD) (a residue of oil distillation) derived from lemon eucalyptus (*Corymbia citriodora*) is the only EPA registered plant-derived active ingredient [17] with DEET-like efficacy [19]. Although not EPA-registered, citronella, cedar, geranium, peppermint and soybean oils are currently marketed due to their safety but their effectiveness have yet to be evaluated [17]. In Australia, the Australian Pesticide and Veterinary Medicines Authority (APVMA) control the sale of insect repellents. APVMA have approved three insect repellents based on *Melaleuca* (tea tree) oil (MOOV insect repellents) [20].

In the last few decades, consumers’ interest in plant-based repellents have increased because they are fully biodegradable, with no ecotoxicology concerns or environmental residues and are considered as ‘safe’ over synthetic repellents [21]. This has led to an increase in plant-based ingredients in insect repellents on the shelves in some countries [22]. Plants synthesise and emit a large variety of volatile organic compounds; floral volatiles serve as attractants for pollinators and volatiles extracted from vegetative parts, especially those release after herbivory, protect plants by deterring herbivores [23]. Essential oils (EOs) obtained from plants through distillation or mechanical methods such as cold pressing are among the best-studied substances. Some plants whose EOs have reported good repellent activity up to 8 h (depends on mosquito species) include amyris, broad-leaved eucalyptus, camphor, catnip, carotin, cedarwood, chamomile, cinnamon oil, juniper, cajeput, *Curcuma longa*, geranium, galbanum, jasmine, litsea, lavender, lemongrass, lemon-scented eucalyptus, narrow-leaved eucalyptus, niaouli, olive, rosemary, sandalwood, soya bean, tagetes and violet [14, 24-27]. EOs contain complex mixtures of volatile organic compounds such as hydrocarbons (terpenes) and oxygenated compounds such as alcohols, aldehydes, esters, ethers, ketones, lactones and phenols [27]. Monoterpenes (α-pinene, cineole, eugenol, limonene, terpiolene, citronellol, citronellal, camphor) and sesquiterpenes (β-caryophyllene) are common constituents of EOs from plants described in the literature as presenting mosquito repellent properties [27]. Most of these EOs vary in the duration of protection against biting mosquitoes with effects lasting for few minutes to several hours [26, 28]. Their active ingredients are highly volatile and they can evaporate quickly from the skin, resulting in brief protection time [14].

In Australia, at least 40 essential oils from native plants, mostly, *Eucalyptus* spp. and *Melaleuca* spp. have been evaluated against mosquitoes, march flies and sand flies [20, 25, 29-31]. Indigenous people from Northern Australia have used smoke produced from burning of blue cypress (*Callistris intratropica* R.T. Baker and H.G. Sm) for generations to repel mosquitoes [32]. Essential oil from the bark and wood of the blue cypress is composed exclusively of sesquiterpene alcohols (guaiol, bulnesol and eudesmol) and terpenes (selinene isomers) [33, 34]. Its antimicrobial activity has been documented, but little is known about its effectiveness to repel insects [20, 33, 35]. The present study was conducted to determine the repellent effect of ABC essential oil and a skin lotion containing the ABC essential oil against mosquitoes. Specifically, the effective doses of the ABC essential oil (active ingredient) and complete protection time of both the active ingredient and the topical formulation (a skin lotion, Aussie Blue Off®) were compared with 20% ethanolic DEET.

## Materials and Methods

### Study design

This study evaluated the repellency of ABC essential oil and a formulated skin lotion containing ABC oil against *Aedes aegypti* in the laboratory and against wild mosquito populations in the field in North Queensland, Australia following World Health Organisation (WHO) guidelines for topical repellent testing [36]. This involved a blinded randomized trial which compared the repellent effect of the test formulations to 20% ethanolic DEET (Merck & Co., Macquarie Park, NSW, Australia) as positive control and 100% laboratory grade ethanol (Merck & Co., Macquarie Park, NSW, Australia) as negative control. Because *Ae. aegypti* bites primarily during the day, the tests were conducted between 9 am and 4 pm.

### Study Participants

Twenty-three volunteers, 13 males and 10 females, having completed an online survey, stating low or no known skin reactions against mosquitoes or ingredients of the products were recruited. The age of the participants ranged from 18 to 65 years. The study was approved by the JCU Human Research Ethics Committee (H8476) and the study participants provided written informed consent prior to repellency testing. To avoid unwanted bias the participants were asked to avoid the use of fragrance and repellent products for 12 h prior to and during testing. During the experiments, the participants were instructed to keep their treated arms away from their clothing and other surfaces.

### Test Formulations

The ABC wood and bark essential oil and a skin lotion containing ABC oil marketed as Aussie Blue Off®, both provided by Australian Blue Cypress Pty Ltd (Northern Territory, Australia) were tested. The ABC oil is referred to hereon, as, active ingredient (AI) and Aussie Blue Off® as topical formulattion. Different concentrations of AI were prepared in laboratory grade ethanol (100 %) prior to each trial while the formulated material was used as supplied.

### Laboratory experiments using arm-in-cage test (AIC)

The laboratory experiments were conducted at the Australian Institute of Tropical Health and Medicine (AITHM), Smithfield, Queensland, Australia from September to October 2021. The mosquitoes used were 3 – 5 days post-eclosion, non-blood fed nulliparous female *Aedes aegypti* (Wb_mel_ strain) from a laboratory-reared colony. The insectary was maintained at 26.0 +/-2.5 °C with a relative humidity of around 75 % +/- 11 %.

#### Effective dose estimation

In order to find the effective dose to prevent mosquito landing or probing, the following (cumulative) doses of ABC oil (AI) were tested: 0.5 %, 1.5 %, 3 %, 5 %, 7.5 % and 10.5 %. The rationale for selecting these doses was that the maximum AI concentration in formulated products was unlikely to exceed 10%. The experiments were repeated 3 times with each volunteer (n = 12) on different days, so that a total of n=36 experiments were conducted. The AI dilutions were applied in increasing concentrations to one of the forearms of each volunteer with a defined amount (e.g., 1 mL per 600 cm^2^) and were exposed to n=50 laboratory-reared *Ae. aegypti* mosquitoes for 30 seconds. The total number of mosquito landings and probings were counted during that period. Results from the triplicate repetitions conducted with each volunteer were averaged. The negative control (ethanol only) (30 s exposure) tests were conducted on each arm prior to testing the AI. The experiments concluded with a negative control on the other, untreated arm.

#### Complete protection time (CPT) estimation

CPT is the number of minutes elapsed between the time of repellent application and the first mosquito landing and/or probing. The AI concentration tested, 20%, was based on the results of effective dose laboratory study (described above) and recommendation in the WHO guidelines [36]. On each volunteer (n = 11), 20% AI was applied to one forearm and 20% DEET was applied to his/her other forearm. After a drying time of 1 min., the treated forearms were exposed to separate cages of n = 200 *Ae. aegypti* mosquitoes for 3 min. The total number of mosquito landings and probings were counted during that period. This was done at 0, 30 and 60 minutes post-application. The formulated product was tested as provided (without any dilution), in the CPT experiments. But the number of mosquitoes per cage was reduced to n=50; WHO guidelines recommended 200-250 mosquitoes per cage [36]. This was due to a substantial biting pressure (over 100 bites within the 3 min exposure period), causing discomfort to the volunteers. As per theAI CPT experiments, the exposure time was 3 minutes and the tests were done at 0, 30 and 60 minutes post-application. WHO guidelines recommend to stop exposure at the time point when the first landing or probing is observed with the candidate product. However, since we never observed complete protection using the AI or formulated product, even directly after application, experiments were only conducted up to 60 min post application.

### Field experiments using human landing catches (HLCs)

The field experiments were conducted in February to April 2022 at Cattana Wetlands in Smithfield, North Queensland, Australia, which is an 80-hectare reserve containing regionally significant forests and a diverse range of bird species and wildlife. This site was chosen due to its high abundance of mosquito species as well as a suitable biting rate. A preliminary assessment of collection sites was performed prior to the trial.

#### Effective dose estimation

The following doses of ABC oil in ethanol were tested in the field: 2.5 %, 5 % and 10 %. The rationale for selecting these doses was that the maximum AI concentration in the formulated products usually does not exceed 5 %. Also, the laboratory results showed that protection increased to around 80 % at the highest AI concentration (10.5% cumulative dose). On each trial day, n=5 volunteers participated and each person was randomly assigned to the 5 solutions including a negative control (ethanol only), a positive control (20% DEET) and the three doses of the AI. Trials were conducted following WHO guidelines [36]. The volunteers were fully covered except for the leg (knee to ankle area) where the test solution was applied. The treated leg was exposed for 10 minutes and any mosquito that landed and/or probed during that period was captured using a hand-held aspirator. The sampling container with mosquitoes were removed from the aspirator. New sampling containers were used for each 10 minutes collection period and each location. Captured mosquitoes were identified in laboratory. The number of mosquitoes that landed but were not captured were also recorded. A single (1-day) test included rotation of each volunteer at random among all (n = 5) collection sites at 10-mins intervals. The experiments were repeated 3 times on different days, so that a total of n=15 experiments were conducted for AI. Results from the triplicate repetitions conducted were averaged.

#### CPT estimation

For CPT, WHO guidelines recommend to stop exposure at the time point when the first landing or probing is observed with the candidate product. Normally, the effective dose that provided 99.9% protection in laboratory studies is used as a guide to establish the dosages for the AI. However, since we never observed a complete protection using the AI in the laboratory, we used a concentration of 5% and 10% to estimate protection against mosquitoes in the field. The rationale for selecting these doses was that the maximum AI concentration in the formulated products usually does not exceed 5%. For ABC oil CPT, n=4 volunteers were recruited per day for testing the two doses of AI and the controls (positive and negative). HLCs were performed as described above. For the topical formation CPT estimation, n=3 volunteers were recruited per day for testing of the formulated material, the positive control and an untreated control. A single (1-day) test comprises rotation of each test volunteer at random among all collection sites (n=3) at 10 minutes intervals. The experiments were repeated 3 times, so that a total of n=9 experiments were conducted for the formulated product. Results from the triplicate repetitions conducted were averaged.

### Data Analysis

Data was analysed using Microsoft Excel 2016 and GraphPad Prism 9.3.1 (GraphPad Inc.). The level of protection (% protection) was calculated as the reduction of mosquito landings and/or probing observed on the treated limb relative to the ethanol-only controls using the formula: Protection (p) = (C-T)/C, where T corresponds to the number of landings/probings on repellent treated arm and C is the average of two control arms (ethanol). Complete protection time is calculated as the number of minutes elapsed between the time of repellent application and the first mosquito landing and/or probing.

## Results

### Effectiveness of Australian blue cypress essential oil (active ingredient) at preventing mosquito bites

Overall, an increasing level of protection with increasing concentrations of AI were observed in both the laboratory (Fig. 1) and field (Fig. 2) experiments. In the laboratory, the median level of protection observed with n=12 volunteers was above 50% for all tested AI concentrations and increased to around 80% at the highest AI concentration (10.5% cumulative dose) (Fig. 1). We observed a large degree of variability in the number of landings and probings with different volunteers both in the control and treatment arms. As such the percent protection calculated was also highly variable, for example, ranging from around 50% to 100 % for a 5% cumulative AI concentration. In the field, the highest dose (10% AI) provided 81% protection (Fig. 2). It should be noted, that 100% protection was rarely achieved at 10% concentration.

**Figure 1.**
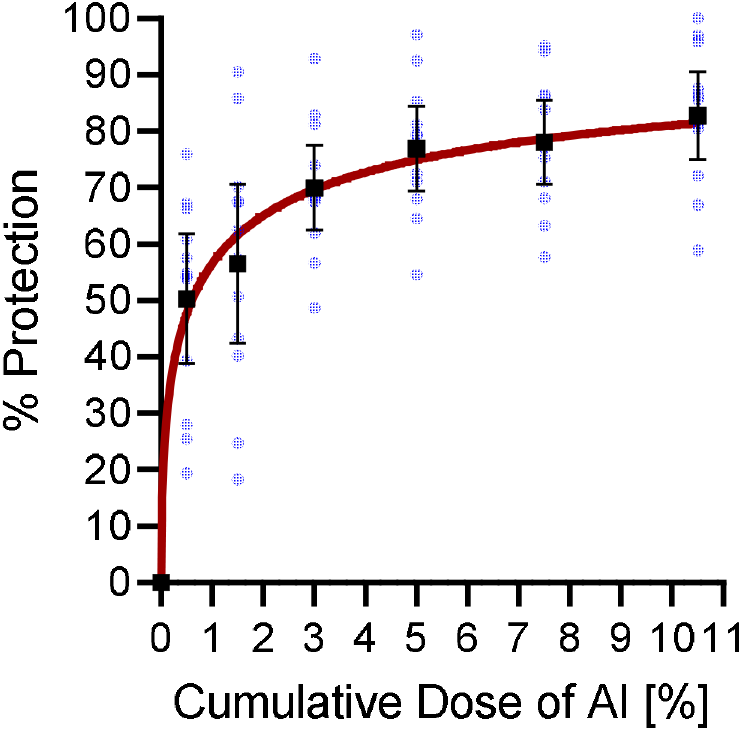
Median level of protection [%, black squares] and 95 % confidence interval for cumulative doses of 0 - 10.5 % of AI (ABC essential oil) from the laboratory. The triplicate data for each of the n=12 participants was averaged (blue dots) prior to the calculation of the medians. The red line is an empiric dose response model.

**Figure 2.**
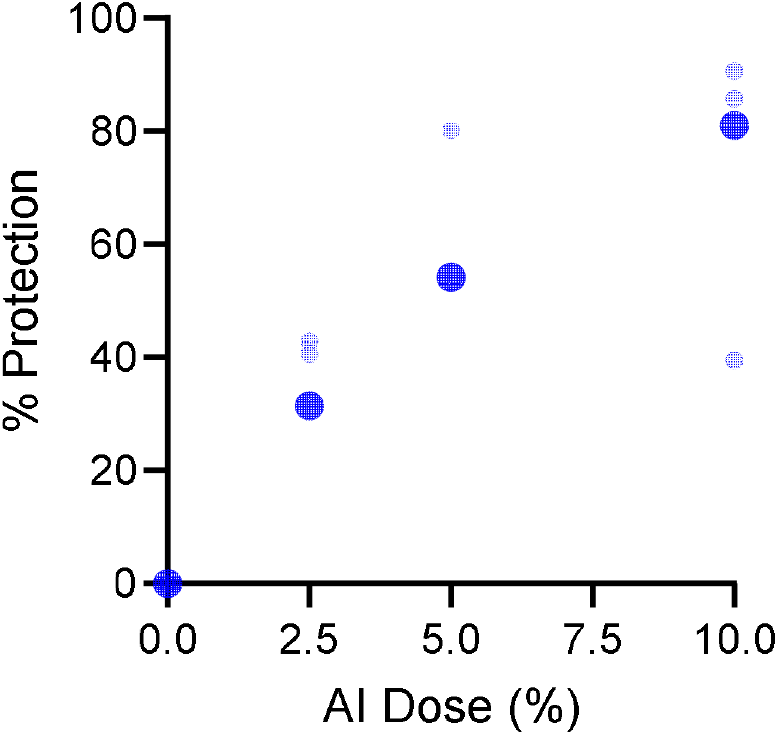
Percent protection for AI (Australia Blue Cypress essential oil) at 2.5, 5.0 and 10.0 % doses from the field. Large blue dots are averages and small, transparent blue dots are results for each replicates for AI. The percent protection for DEET (not shown) was always at 100 %.

### Complete protection time

In the arm-in-cage tests, CPT for either the 20% ABC essential oil or formulated skin lotion were <1 min because mosquito landing and probing were recorded at the exposure immediately after application. The percent protection and the absolute number of mosquito landings and probings observed in the 3 min exposure over time afforded by DEET, AI observed in the 11 volunteers, and formulated skin lotion observed in the 8 volunteers is shown in Fig. 3. A large degree of variability was observed in the number of landings and probings among volunteers with both the AI and the formulated skin lotion. Generally, DEET (as the positive control) prevented probing to 100% but allowed for sporadic landing during the one-hour observation period. This is because the chemorepellency of DEET is mediated by *Ae. aegypti* feet upon alighting on the skin [37]. There were instances of probing at 60 min after treatment with DEET in the case of two volunteers in the formulated product CPT trials but we attribute this to the volunteer not following instructions i.e. rubbing off the repellent during the trial rather than a failure of DEET. The percent protection afforded by 20% ABC diluted in ethanol against 200 *Ae. aegypti* mosquitoes was 81% and decreased to 73% after 1h (Fig.e 3A and C). This was statistically significantly less protection than the DEET (p<0.0001) and meant that with 200 mosquitoes per cage, volunteers still received more than an average of 50 landings and/or probings during the 3 min exposure period. With the formulated skin lotion, we observed a slightly higher initial % protection values of about 85% which then decreased to around 75% at 1h post-application (Fig. 3B and D). With 50 mosquitoes per cage that meant that volunteers still received about 10-20 bites in the 3 mins exposure period.

**Figure 3.**
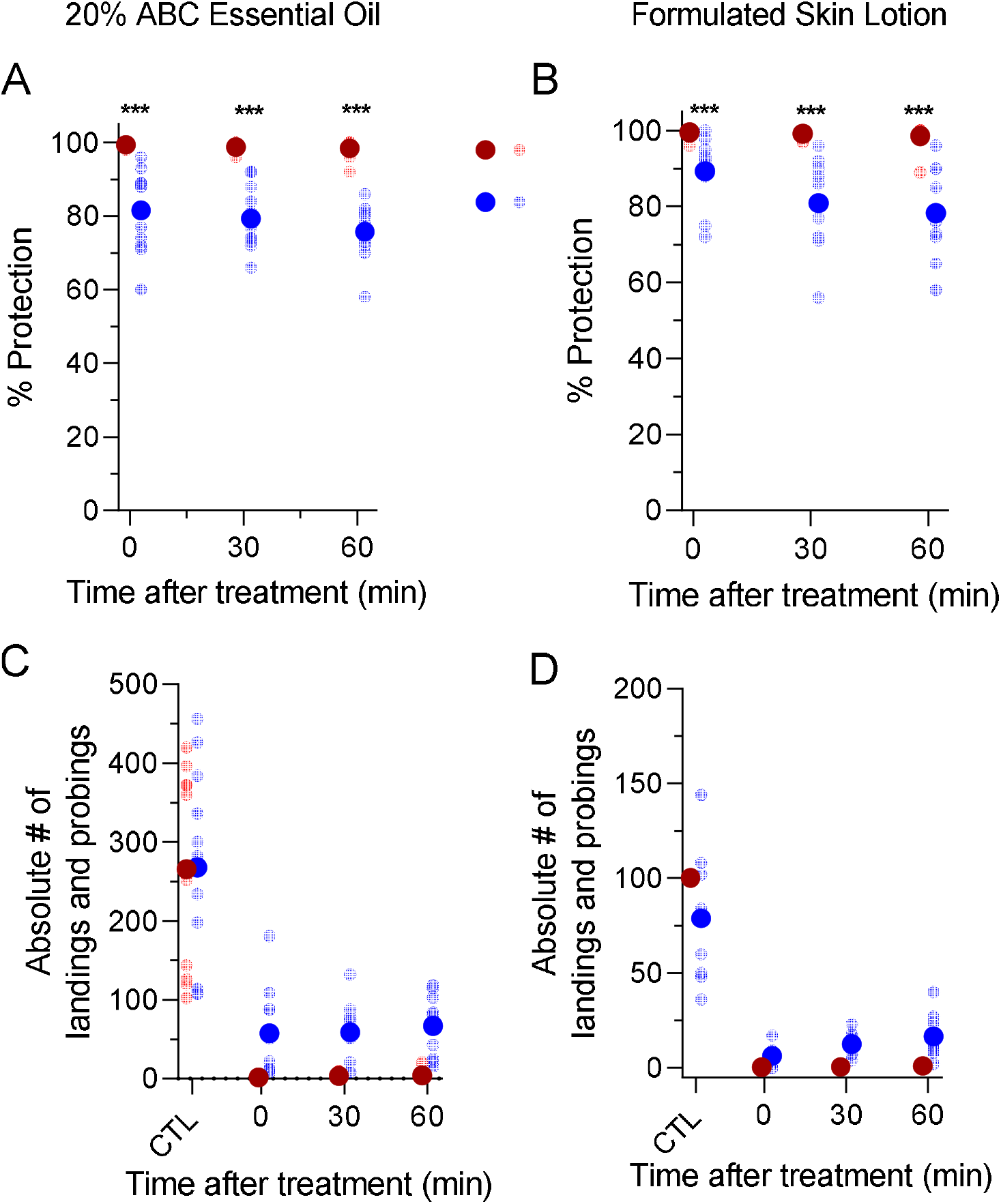
Complete protection time experiments with 20% ABC essential oil and ABC formulated skin lotion from the laboratory AIC tests. Panels A and B show the % protection (i.e., the number of landings or probings relative to the negative control) over time. Panels C and D show the absolute number of bites received before (control, CTL) and after treatment. Large dots are averages and small, transparent dots are results for each replicate (volunteer). Red is DEET and blue is for candidate product. *** indicates statistical significance (p<0.0001) between DEET and candidate product (Mann-Whitney U test)

In the field, 10% ABC oil induced 93% protection, which decreased to 56% after 50 min while 5% ABC oil induced 75% protection, which decreased to 50% protection after 50 min (Fig. 4A – B). Similarly, protection decreased from 63% to 50% for the skin lotion (Fig. 4C). In comparison, DEET maintained a 100% protection throughout the testing period of 50 mins and there were no landing/probing observed in volunteers who had applied DEET. For comparison, we also show the absolute number of mosquito landings and/or probings observed in the 10 min exposure period for ethanol-only (negative control), DEET, AI and the formulated skin lotion in Fig. 4 D – F.

**Figure 4.**
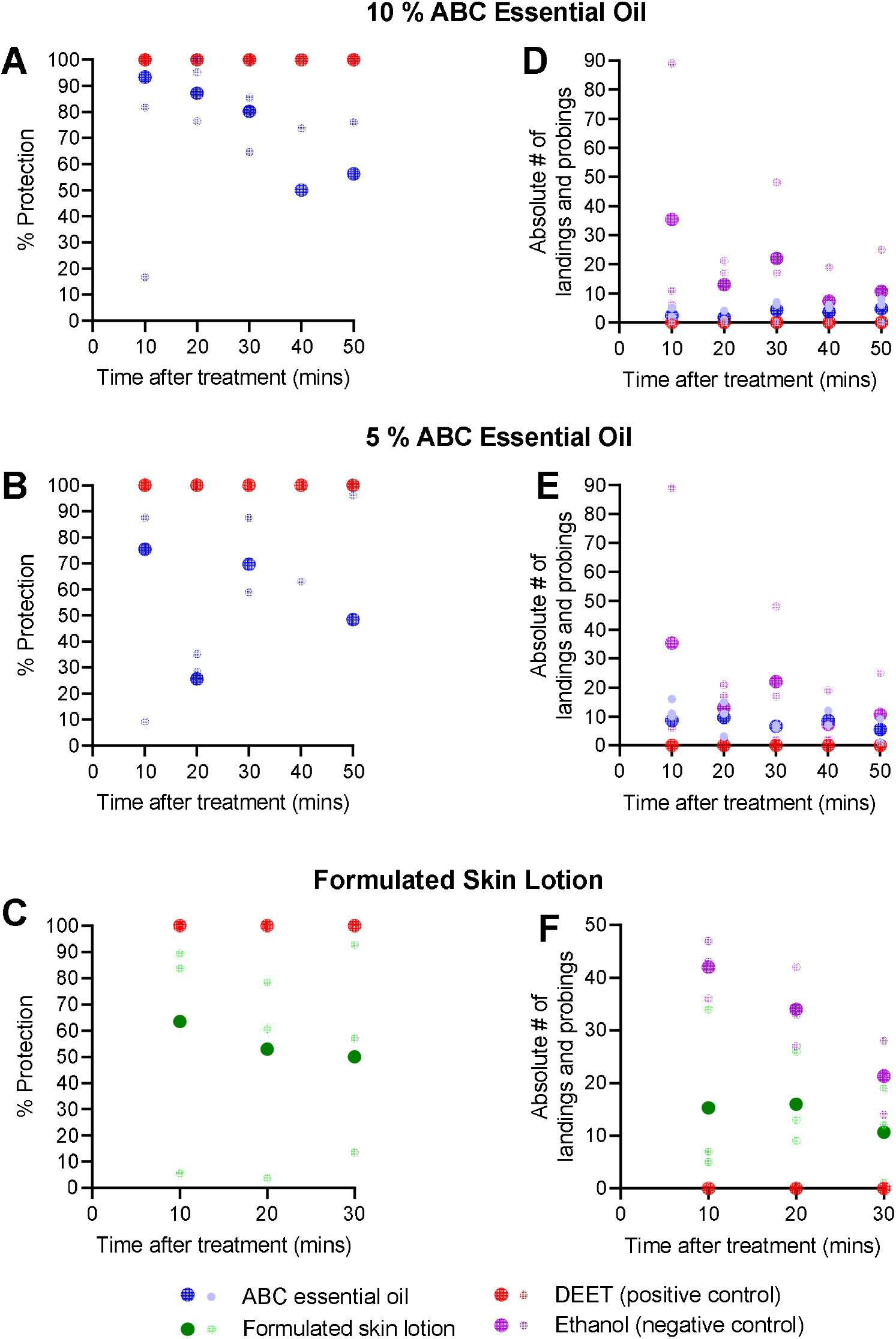
Complete protection time experiments with ABC essential oil and formulated skin lotion in the field HLC trials. Panels A, B and C show the % protection (i.e., the number of landings or probings relative to the negative control) over time. Panels D, E and F show the absolute number of landings/probings received by volunteers who applied ethanol (negative control), candidate products and DEET (positive control). Large dots are averages and small, transparent dots are results for each replicate (volunteer). Blue is AI (Australian blue cypress essential oil), Green is formulated skin lotion (Aussie Blue Off Lotion), red is DEET (positive control) and purple is ethanol (negative control).

### Mosquito species composition

Altogether n=640 mosquitoes belonging to eight species were caught in the field on legs treated with ABC oil and the formulated skin lotion. The most common species were *Verralina carmenti* (63% and 59%) and *Ve. lineata* (30% and 29%) for ABC oil and formulated skin lotion, respectively (Fig. 5). Others caught in low numbers (≤ 7%) included *Aedes. vigilax, Ve. funera, Ae. notoscriptus, Ae. kochi, Culex annulirostris* and *Coquillettidia crassipes*. The details of collected mosquito species and numbers are in Supplementary Table S1 (https://doi.org/10.6084/m9.figshare.21985679.v1).

**Figure 5.**
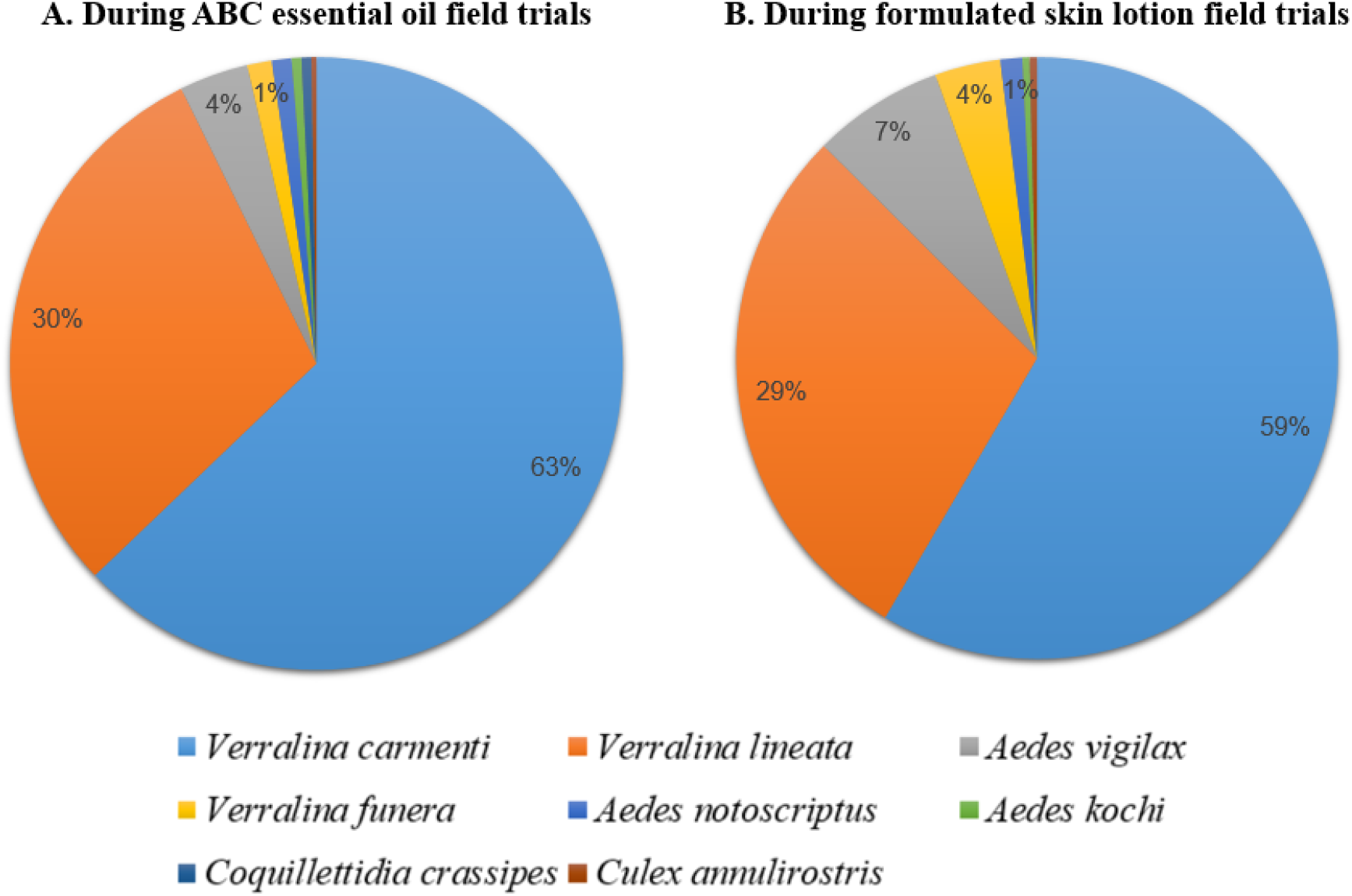
Overall species distribution of mosquito species caught using HLC method at Cattana Wetlands, Smithfield, Queensland from February to April 2022. Mosquito species from treated legs with ethanol-only (negative control) and candidate products are combined. Panel A shows percentage of species caught ABC essential oil field trials while Panel B shows percentage of species caught during formulated skin lotion trials.

## Discussion

The presence of sesquiterpenoid and terpene indicate that ABC products could possess repellent activity (Fig. 1 and 2). *C. intratropica* essential oil is composed mainly of guaiol (26%), bulnesol (16%), eudesmol isomers (9.5%) and selinene isomers (19 - 22%) as well as unique lactones (<1%) like callitrin, callistrisin and columellarin [34, 38]. However, it should be noted that the composition of bioactive compounds depends on the collection site and the *Callistris* species. For example, the guaiol in wood essential oil of *C. intratropica* (blue cypress) is 17 – 21% and in both *C. columellaris* (white cypress) and *C. endlicheri* (black cypress), is 50%.

The blue cypress essential oil diluted in ethanol (5%, 10% and 20%) and the formulated skin lotion provided moderate protection, preventing about 70 - 90% of landings and probings directly after application compared to DEET (Fig. 3 and 4). In addition, the complete protection time was short (less than a minute) for both the 20% ABC oil diluted in ethanol and a formulated skin lotion against *Ae. agypti* (Fig. 3). We searched the major databases, Pubmed, Scopus and Google Scholar, with keywords ‘blue cypress’, ‘*Callitris intratropica’*, ‘essential oil’, ‘insect’, ‘repellent’ for relevant papers but very little documentation was available. In addition, due to the variability on methodologies found in the literature to assess repellency, it makes it difficult to compare our results with reported studies. Consequently, we will compare our results to studies whose methods were similar to WHO guidelines and the bioactive compounds in essential oils of plants that have repellent activity.

Greive et al., 2010 also reported short protection times from Australian *C. collumelaris* (5% wood EO in cream base) and *C. glaucophyla* (5% wood EO in cream base) against *Ae. aegypti* but they did not report information on the percent protection. In a laboratory study, Trongtokit, al., 2005 compared the repellency of some of the common essential oils against 250 mosquitoes with an exposure time of 1 min (ours was 3 min). Of the three most effective oils, (clove, citronella and patchouli), 10% clove oil provided 100% protection against *Ae. aegypti* up for 30 mins, while all three provided up to 2 h of protection against *Cx. quinquefasciatus* and *An. dirus* [28]. The major constituents of clove oil are eugenol, eugenol acetate and beta-caryophyllene [28].

Another study showed that mixtures of Australian *C. glaucophylla* with other botanicals resulted in 100% mortality against *Cx. annulirostris* larvae and affected emergence of *Ae. aegypti* adults [39]. In addition, sandalwood essential oil, which contained a higher percentage of guaiol (43.8%), appeared to have the most effective larvicidal activity on *Ae. aegypti, Ae. albopictus* and *Cx. pipiens* compared to cinnamon oil, lemon eucalyptus oil and turmeric oil [40]. Further studies on the effect of *C. intratropica* essential oil on mosquito larvae are needed.

Repellents that have low volatility; for example, PMD, a monoterpene, derived from lemon-scented eucalyptus (*Corymbia citriodora*) does not tend to evaporate rapidly after application on the skin, resulting in longer protection time [26, 41]. In another study, essential oils of clove, citronella grass and lemongrass diluted in coconut oil or olive oil, induced protection up to 96 mins for *Ae. aegpyti* and 165 mins for *Cx. quinquefasciatus* [42]. To extend the duration of repellency, several studies have added 5% vanillin to essential oils of plants such as turmeric, citronella grass, hairy basil and eucalyptus oil, which repelled mosquitoes for up to eight hours [25, 26, 43, 44]. The essential oil of Australian *C. intratropica* is highly volatile resulting in brief protection times, which maybe improved with addition of other botanicals as synergists, such as 5% vanillin.

Here, we showed that 20% ethanolic DEET maintained a 100% protection during the entire study duration in both AIC (60 min) and HLC experiments (30 min and 50 min) (Fig. 3 and 4). Studies have shown that DEET at concentration of 20% or more provided the best efficacy against *Aedes* species with protection up to 10 h whereas at 4 – 15%, protection lasted 1 and 7.5 h [26]. DEET has at least two molecular targets for mosquitoes: odorant receptors (ORs) mediate repellency at a distance and chemoreceptors mediate repellency upon contact [45]. The underlying mechanisms by which DEET works is unclear. It is thought to work by blocking the insect’s odorant receptors (olfactory receptor, ORx) which detect l-octen-3-ol, found in human breath and sweat [46]. Interestingly, previous studies have found that essential oils containing oxygenated compounds have a higher repellent activity than DEET against *Ae. aegypti* and *An. gambiae* [47, 48].

## Conclusions

The results of this study indicate that *C. intratropica* essential oil offers some protection against biting mosquitoes. Adding a botanical synergist may offer commercial potential as a short-period repellent or under conditions of low mosquito abundance. However, it is important that public health messages continue to emphasize the greater effectiveness of DEET-based repellents in areas with risks of mosquito-borne disease.

## Data Availability

All data produced in the present study are available upon reasonable request to the authors.

## Acknowledgements

We thank Australian Blue Cypress Pty Ltd. for their partnership and for providing the test formulations. We are grateful to the study participants for their great collaboration in the laboratory and field experiments. We acknowledge the Cairns Regional Council for granting permissions to conduct field trials at Cattana Wetlands in Smithfield. The work received funding from Innovation Connections Researcher Placement grant (ICG001564) and Australian Blue Cypress Pty Ltd.

## Author Contribution

SK conceived the study; BA conducted online surveys and initiated recruitment of volunteers; BA, MK and SK designed the study and conducted the laboratory experiments; MK, SK and MT conducted the field experiments; MT reared *Aedes aegypti* mosquitoes and identified collected mosquitoes. MK and SK analyzed the laboratory and field data. MK wrote the first draft. All authors read and approved the final manuscript.

## Additional Information

### Data availability

The datasets generated during and/or analysed during the current study are available from the corresponding author on reasonable request.

